# The Efficacy of Nurse-Performed Ultrasound Guidance Compared with the Conventional Cannulation Technique in Patients with Difficult Peripheral Intravenous Access: A Systematic Review

**DOI:** 10.1101/2022.06.01.22275477

**Authors:** Lisa Maria Anderssen, August Gabriel Wang, Anna Sofía Fjallheim

## Abstract

**Objective:** The aim of this review is to highlight the efficacy of nurse-performed ultrasound guidance compared with the conventional cannulation technique in patients with difficult peripheral intravenous access.

**Design:** A systematic litterature review.

**Data sources:** The CINAHL and PubMed databases were searched for articles from the period 2011-2021.

**Method:** The following search words were used: peripheral intravenous AND ultrasonography OR ultrasound guided AND catheterization, peripheral/methods. The keyword catheterization, peripheral/methods was found via MeSH Terms (Medical Subject Headings) which PubMed recommended as keyword within the intervention of the conventional cannulation technique.

**Results:** 2 out of 3 articles prove that success rate on the first attempt (primary outcome) was significantly higher in the nurse-performed ultrasound-guided technique compared with the conventional palpation technique. The results of the secondary outcomes; time consumption, complications, patient satisfaction and nurse satisfaction between the two groups proved to be heterogeneous.

**Conclusion:** Nurse-performed ultrasound guidance in hospital wards increases the success rate in patients with difficult peripheral intravenous access.

## Introduction

Peripheral intravenous (IV) cannulation is an invasive lifesaving procedure performed in patients worldwide. Each day peripheral intravenous catheters (PIVC) are placed in 60-90% of patients admitted to hospital.^1,2^ Fluids, blood products, medications and nutrients can all be given via the IV route and up to 80% of hospitalized patients receive infusion therapy.^3,4^ In previous years, only doctors were qualified to insert PIVCs. But in the early 1970s a revolution occurred when the first nurse, Ada Plumer, was permitted to insert the first PIVC. This led to the establishment of the Intravenous Nurses Society in 1973.^5^ The specialty in IV access within the nursing profession has expanded greatly since then and today nurses administer PIVCs as a part of their clinical expertise. A PIVC is usually inserted in veins on the upper extremity, but can be placed in other sites of the body. Most PIVCs are inserted using the conventional technique (CT), which requires visual inspection and palpation of the patient’s veins.

Obtaining an IV access is not always a straightforward procedure, especially in patients with a history of difficult IV access (DIVA). Up to one-third of all patients have DIVA.^6,7^ DIVA is often due to lack of visual or palpable veins caused by different factors such as obesity, edema, smaller veins, vascular pathology, intravenous drug abuse, fluid status, chronic illness, burn injuries, hypovolemia, malnutrition and chemotherapy. DIVA is also common in the paediatric and elderly patient population.^6-12^ Patients with DIVA often have to undergo multiple failed insertion attempts by different hospital staff. Failed insertion attempts lead to vessel trauma that increases the risk of complications such as phlebitis, infiltration, extravasation, occlusion, dislodgment and bloodstream infection.^1,13^ Unsuccessful attempts result in painful patient experiences and discomfort and blood draw and laboratory test results are delayed causing further delay in diagnoses and medical treatment. Furthermore, the patients experience increased pain associated with multiple IV attempts.^7,14^ Patients who fail PIVC access are often rescued by placement of central venous catheters (CVC), which is a far more invasive procedure, time consuming, expensive and may lead to serious complications such as pneumothorax.^9,15^

Ultrasound guidance (UG) has been recommended for the placement of PIVCs in patients with DIVA to reduce needle insertion attempts and the total procedure time. UG has also been recommended as an effective alternative to CVC insertion in patients with DIVA.^16^ Ullman and colleagues first described an ultrasound-guided technique for the placement of CVC in 1978.^17^ The first study of UG on peripheral veins was conducted in 1999 by Keyes and colleagues, which concluded UG IV catheterization to be more successful than the CT.^18^ The use of UG to facilitate PIVC access in patients with DIVA has since then expanded greatly.

UG can be performed through different techniques. The Dynamic Needle Tip Positioning (DNTP) technique is an ultrasound technique where the beam intersects the target vessel in the cross-sectional plane displaying a short-axis view of the vessel. What makes the DNTP technique so special is that it has a significantly higher success rate compared to the longitudinal long-axis view (97% vs. 81%). Furthermore, the DNTP technique has the remarkable possibility to track the needle tip with high accuracy in real time visualization, placing the needle tip in the centre of even small vessels.^19^

There is little information with regards to how many systematic reviews exist that compare nurse-performed UG with the conventional cannulation technique in patients with DIVA.

The aim of this study was to systematically review the results of randomized controlled trials comparing UG with the conventional cannulation technique administered by nurses on clinical wards. This was done in order to highlight the efficacy of clinical nurse-performed UG when used on hospitalized patients with DIVA. The primary outcome was success rate (placement of a functional PIVC) on the first attempt. The secondary outcomes measured time consumption (total time to achieve functional vascular access), complications, and patient satisfaction and nurse satisfaction. The hypothesis is that there is efficacy in nurse-performed UG.

## Methods

This systematic review was conducted in compliance with the Preferred Reporting Items for Systematic Review and Meta-Analysis (PRISMA) guidelines.^20^

### Search Strategy

Databases of peer-reviewed literature were systematically searched and originally performed in April/May 2021. The PubMed and CINAHL databases were searched for data spanning from January 2011 until January 2021. These databases were chosen in this review because they comprise aspects of health sciences, medicine and nursing. In addition, these databases are internationally recognized. The keywords were found through pilot searching and were listed in this order: peripheral intravenous AND ultrasonography OR ultrasound guided AND catheterization, peripheral/methods. The keywords *catheterization, peripheral/methods* were found via MeSH Terms (Medical Subject Headings), which PubMed recommended as the keywords within the intervention of the conventional cannulation technique.

### Study Selection and Data Extraction

Eligibility criteria of the studies included in this systematic review were defined beforehand, but no formal review protocol was recorded. Studies were eligible for inclusion in this review if they met the following criteria: (1) randomized controlled trials, (2) nurse-performed IV cannulation, (3) adult patients requiring PIVC access, (4) patients randomized to UG versus the conventional technique for the placement of PIVC, (5) patients with DIVA, (6) English language, (7) peer-reviewed research articles, (8) all countries included. Studies were excluded for the following reasons: (1) IV insertion of other devices e.g., arterial catheters, central venous catheters, etc., (2) children, (3) if the UG technique was not compared with the conventional palpation technique. Articles were reviewed by title, by abstracts and by full-texts. The flowchart of the study selection procedure is presented in Figure 1. From the database search, 5 randomized controlled trials (RCT) came forth through the process of title screening and abstract screening. After a thorough full-text screening only 3 RCT studies were eligible.

**Fig 1.**
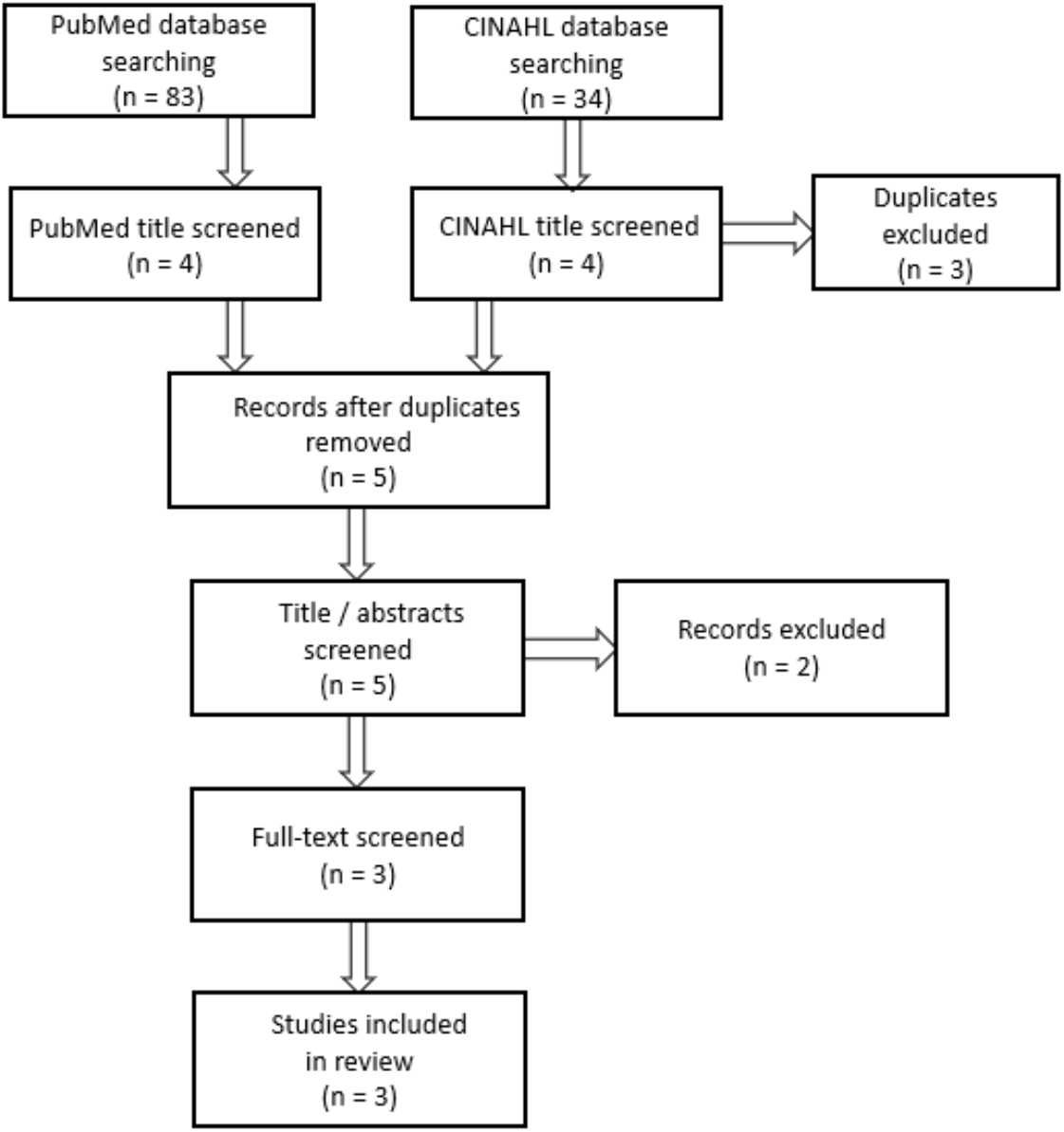
Flowchart of Study Selection.

### Outcome Measures

The primary outcome was success rate on the first attempt. The secondary outcomes measured time consumption to achieve functional vascular access, complications, patient satisfaction and nurse satisfaction.

### Statistical Analyses

A *P*-value < .05 was considered statistically significant.

## Results

After the articles had been systematically reviewed, including removal of duplicates, title, abstract and full-text screening, a total of 3 RCTs were selected that met the inclusion and exclusion criteria (see Table 1).

**Table 1.** Overview and Results.

| First author and year | Design | Setting | Participants | Sample Size | Outcomes Measured | Outcome |
| --- | --- | --- | --- | --- | --- | --- |
| Bahl 2016<sup>21</sup> | RCT | ED in USA | ED patients with DIVA | 124 (63 patients in the UG group and 61 in the CT group) | Success rate on the first attempt, time consumption and IV attempts per subject | Success rate 76% vs. 56% ($p = .02$)<br>Time consumption 20.7 min vs. 15.8 min ($p = .75$)<br>IV attempts per subject 1.52 vs. 1.71 ($p = .63$) |
| Bridey 2018<sup>22</sup> | RCT | ICU in France | ICU patients with DIVA | 114 (57 patients in each group) | Success rate on the first attempt, duration of the PIVC, PIVC diameter, patient satisfaction and nurse satisfaction | Success rate 41% vs. 33% ($p = .886$)<br>Duration of the PIVC 3 days vs. 3 days ($p = .719$)<br>PIVC diameter 18 gauge vs. 21 gauge ($p = .059$)<br>Extravasation 34% vs. 18% ($p = .094$)<br>Patient satisfaction 8 vs. 8 ($p = .543$)<br>Nurse satisfaction:<br>Clinical setting ($p < .001$)<br>Time-saving ($p = .018$)<br>PIVC quality ($p = .017$) |
| Nishizawa 2020<sup>23</sup> | RCT | ICU in Japan | ICU patients with DIVA | 60 (30 patients in each group) | Success rate on the first attempt, success rate after two attempts and complications after PIVC placement | Success rate 1. attempt 70% vs. 40% ($p = .0379$)<br>Success rate 2. attempt 73,3% vs. 46,6% ($p = .0651$)<br>Extravasation 13,6% vs. 28,6% ($p = .394$) |

Abbreviations: CT, conventional technique; DIVA, difficult intravenous access; ED, emergency department; ICU, intensive care unit; IV, intravenous; PIVC, peripheral intravenous catheters; RCT, randomized controlled trial; UG, ultrasound guidance.
<sup>a</sup>Significant at $P < .05$ .

The first article is an American RCT from 2016.^21^ This study compares nurse-performed UG with the CT in DIVA patients admitted to the emergency department (ED). The results showed that there was a statistically significant difference in the success rate between the two groups (*P =* .02). The success rate for nurse-performed IV placement was 76% in the UG group, compared with 56% in the CT group. The mean time it took to place a successful IV access was 20.7 minutes in the UG group versus 15.8 minutes in the CT group and showed no statistical difference regarding time consumption (*P* = .75).

The next article is a French RCT from 2018.^22^ This study compares nurse-performed UG with the CT in DIVA patients admitted to the intensive care unit (ICU). The results showed no statistical difference regarding success rate on the first attempt, 41% (UG) versus 33% (CT) (*P* = .886). Patient satisfaction did not significantly differ between the two groups (*P* = .543). Extravasation tended to be more frequent in the UG group than the CT group (34% vs. 18%, *P* = .094). The study showed that the UG technique was more suitably adapted and was significantly better regarding nurse satisfaction measured as clinical setting (*P* < .001), time-saving (*P* = .018) and PIVC quality (*P* = .017).

The third article is a Japanese RCT from 2020.^23^ This study compares nurse-performed UG with the CT in DIVA patients admitted to the ICU. The success rate on the first attempt was significantly higher in the UG group compared with the CT group (70% vs. 40%, *P* = .0379). Extravasation occurred in 13,6% of patients in the UG group compared with 28,6% of patients in the CT group, but did not significantly differ between the two groups (*P* = .394).

## Discussion

This review investigates the efficacy of nurse-performed UG in patients with DIVA admitted to hospital. Studies already exist concerning the efficacy of UG in patients with DIVA, however, other professionals such as doctors and emergency technicians perform this procedure. Two out of the three RCTs included in this study proved that nurse-performed UG significantly increases the success rate of intravenous cannulation when compared with the CT.^21,23^ The third RCT article did not have a significant difference between the two methods.^22^ Regarding the primary outcome, it seems evident that the use of UG performed by expertly trained nurses increases the success rate already on the first attempt in patients with DIVA.

A systematic review and meta-analysis from 2018, van Loon et al. included 8 articles that showed a significant difference in favour of UG (*P* = .003).^8^ Another systematic review and meta-analysis from 2013, Egan et al. also showed a significant efficacy in favour of UG (*P* = .008).^9^ A very large RCT from 2016 from an emergency department proved significant favour in the use of UG in 1189 patients with DIVA.^24^ Their success rate in patients with DIVA was 81,6% (UG-group) versus 35,1% (CT-group).

A Danish prospective, descriptive study with a historical control showed that after implementing systematic training of nurses in the use of UG, the use of CVCs was significantly reduced from 45,8% to 13,2% in patients receiving apheresis treatments (*P* < .001).^15^ Besides this, their first attempt success rate in nurse-performed UG was 96,5%. These nurses were trained in performing the DNTP technique.^19^ A retrospective observational study was performed on the same Danish hospital ward three years later and showed that the use of nurse-performed UG for the placement of PIVCs continued to have a positive efficacy on the patients where 97,4% of the apheresis procedures were accomplished via peripheral venous access.^25^ These studies show that there is strong evidence of the positive efficacy in the use of UG in patients with DIVA.

The results of the secondary outcomes were very heterogeneous with regard to time consumption, complications, and patient satisfaction and nurse satisfaction. Furthermore, some of these factors, like nurse satisfaction, have been little investigated. The French RCT study showed that there was significantly higher nurse satisfaction in the UG group compared with the conventional method.^22^ On the other hand, no significant difference was found between the groups with regard to patient satisfaction.

Van Loon and colleagues found in 4 of their studies significantly better patient satisfaction in the UG group (*P* < .001).^8^ A cohort study from 2009, Bauman et al. also proved significantly higher patient satisfaction in favour of UG (*P* < .0001).^26^

The American RCT study did not have any significant difference in the total time it took to achieve a successful IV access.^21^ Bahl and colleagues state that there were factors that could have contributed to the lack of difference in the total time to IV placement, which was the possibility to use ultrasonography as a rescue option in the control group.^21^ There would be a far greater difference in the time to IV placement if the investigators of the study had withheld the use of UG in the group using the traditional palpation technique. The investigators believed it to be unethical to withhold the rescue option of nurse-placed ultrasound-guided IV cannulation in the control group and this could have acted as a confounding factor with regard to the difference in time consumption. An RCT study from 2020, Skulec et al. with 300 participants showed that there was a significant shorter procedure time in the intervention group.^27^

The French and the Japanese RCTs measured the incidence of extravasation complications occurring in each of the two groups.^22,23^ The French study, Bridey et al. showed that the incidence of extravasation was more frequent in the UG group compared with the CT group but with no significant difference.^22^ Bridey et al. state that the reason for more frequency of extravasation in the UG group was due to the use of catheters with insufficient length for the depth of the punctured veins. On the contrary, the Japanese study Nishizawa et al. had fewer occurrences of extravasation in the UG group compared with the CT group, but did not have any significant difference either.^23^ Nishizawa et al. refer to the French RCT and explain the importance of using the right catheter length.^22^ The basilic or cephalic veins, which anatomically run deeper, are most often selected when using the UG procedure. Nishizawa et al. stated in their study that the use of longer catheters in the UG group might have led to the few incidences of extravasation.^23^

Egan and colleagues did not investigate the incidence of complications in their systematic review and meta-analysis.^9^ Van Loon et al. did, on the other hand, find two studies investigating the complications, such as the incidence of infiltration, which is similar to extravasation.^8^ But here they did not find any significant difference between the methods. Skulec et al. did investigate the complication of extravasation, but did not find any significant difference either.^27^ Bauman et al. on the other hand, had fewer complications in the UG group compared to the traditional method (41,5% vs. 64,7%).^26^ UG performed by trained physicians has been shown to have fewer complications in patients with DIVA.^18,28^ An observational study from 2004, Brannam et al., investigating nurse-performed UG in the emergency department also showed to have fewer complications.^29^

There are international recommendations on ultrasound-guided vascular access in patients with DIVA. An international conference in 2012 (International Consensus Conference on Ultrasound Vascular Access) has formulated an evidence-based recommendation where UG is recommended in patients with DIVA.^30^ The American Association for Vascular Access (AVA) recommends use of real-time UG (DNTP technique). AVA confirms that the use of the DNTP technique for vascular access is recommended by multiple organizations, associations, guidelines and standards.^2^ AVA states that when the DNTP technique is performed by trained, competent clinicians, it has been shown to reduce multiple cannulation attempts and decrease complications and, as a result, has improved patient safety, patient satisfaction and cost reduction. Furthermore, they expect that the use of UG will continue and the overall usage of these evidence-based technologies will expand in the coming years. The American Society of Hospital Medicine published recommendations in 2019 where they recommend the use of real-time UG (DNTP) in patients with DIVA. They also state that the use of ultrasound by nurses for PIVC placement has been shown to reduce the time in obtaining venous access, improve patient satisfaction, and reduce the need for physician intervention.^16^

A limitation of this review is that it only includes 3 RCT studies which all take place in single departments. Furthermore, the studies take place in 3 different countries, which can be difficult to compare. However, they all investigate the efficacy of nurse-performed UG on adult patients with DIVA. The homogeneity of this study is therefore quite strong. Another strength of this study is that the authors only used qualified databases, PubMed and CINAHL. However other databases may have given different search results. Also the use of MeSH terms may have narrowed the search results.

Databases of peer-reviewed literature were systematically searched and originally performed in April/May 2021, and at that time there was no existing systematic review that exclusively focused on the success rate of nurse-performed UG on adult patients with DIVA. It was not until recently in March 2022 that the Journal of Emergency Nursing published the first systematic review and meta-analysis on the efficacy of nurse-performed UG versus the traditional peripheral venous access.^31^ The researchers of this study, Tran et al., state that they focused on this specific nurse-performed intervention since previous meta-analysis did not address the important question of whether nurses can perform ultrasound-guided peripheral venous cannulation effectively. Tran et al.’s study had the same primary outcome as our study, which was the success rate on the first attempt. Tran et al. also had other outcomes such as procedure length (length in minutes) and patients’ satisfaction. Their results showed that UG performed by nurses was associated with a significantly increased success rate on the first attempt compared with the conventional landmark-based practices (*P* < .001).^31^ Tran et al. did not find any statistical difference with regards to patient satisfaction and procedure length. They state that few conclusions can be drawn from their analysis aside from the need for further investigation regarding their secondary outcomes, which are PIVC attempts, procedure length and patient satisfaction. Furthermore, they refer to the same three RCT studies, which are included in this study and specify that these studies are the only RCT’s that reported complications or adverse events.^21-23^ Given that our study investigated the occurrence of complications, we can state on behalf of the few existing RCT studies on this subject, that there is a need for more studies investigating this important topic. Tran et al. mention Nishizawa et al. to be the most recent RCT study existing, which gives us a clear indication that our study contains the three newest RCT studies on this topic. Three out of the seven RCT’s included in Tran et al.’s study were assessed as having low risk of bias and these same three RCT studies are also included in our study.^21-23^

The authors recommend the use of UG directly on patients with DIVA on the first cannulation attempt as a standard method and not as a rescue method.^21^ This recommendation includes of course the necessity of adequate training of nurses in the UG technique and access to the ultrasound technology on the wards. As an example, there already exists a Danish e-learning program, which proved to be successful.^15,25^ The authors recommend for future research, RCT studies of nurse-performed UG with more participants that also include technical factors such as the use of DNTP technique and focus on catheter length and size.22,23

## Conclusion

This systematic review demonstrates that nurse-performed UG on adult patients with DIVA increases the success rate compared to the CT. However, this necessitates adequate nurse training and access to the UG technology on the hospital wards.

## Data Availability

All data produced are available online at pubmed and CINAHL

## Author contributions

Lisa Maria Anderssen, RN, was the first author of this study. Ms. Anderssen was responsible for the conception and design of the study and acquired the data and participated in the analysis and interpretation of data and drafting the article. She is a recently graduated nurse from the Danish Deaconess Foundation. Her interests are in research of nursing care.

August Gabriel Wang, MD, DMSc, Professor, was the coauthor of this study. Dr. Wang participated in the analysis and interpretation of data and drafting the article. Dr. Wang is affiliated as a professor to the University of the Faroe Islands and is currently working at the Copenhagen University Hospital. He has 40 years of research experience in suicidology, genetics and patient care and has made several publications.

Anna Sofía Fjallheim, RN, MScN, PhD. Fjallheim was responsible for thoroughly reading through the manuscript and making additional comments. Fjallheim is a teaching lecturer in nursing at the Faculty of Health Sciences at the University of the Faroe Islands.

## Acknowledgements

The authors would like to give special thanks to the Danish Deaconess Foundation for believing in this review from the very beginning and encouraging its publication, which is based on Ms. Anderssen’s bachelor dissertation. Thanks to Mourits Joensen for technical assistance of this project. Thanks to the University of the Faroe Islands for their affiliation with this project.

## References

1. Helm RE, Klausner JD, Klemperer JD, Flint LM, Huang E. Accepted but Unacceptable: Peripheral IV Catheter Failure. J Infus Nurs. 2015;38(3):189–203. doi:10.1097/NAN.0000000000000100

2. Thompson J, Garret JH. Guidance Document. Transducer disinfection for evaluation and insertion of peripheral and central catheters for vascular access teams and clinicians. JAVA. 2018;23(3):141–146. doi:10.1016/j.java.2018.08.001

3. Tripathi S, Kaushik V, Singh V. Peripheral IVs: Factors Affecting Complications and Patency – A Randomized Controlled Trial. J Infus Nurs. 2008;31(3):182–188. doi:10.1097/01.NAN.0000317704.03415.b9

4. Vitto MJ, Myers M, Vitto CM, Evans DP. Perceived Difficulty and Success Rate of Standard Versus Ultrasound-Guided Peripheral Intravenous Cannulation in a Novice Study Group. A Randomized Crossover Trial. J Ultrasound Med. 2016;35(5):895–898. doi:10.7863/ultra.15.06057

5. Rivera AM, Strauss KW, Zundert AV, Mortier E. The history of peripheral intravenous catheters: How little plastic tubes revolutionized medicine. Acta Anaesthesiol Belg. 2005;56(3):271–282.

6. Blanco P. Ultrasound-guided peripheral venous cannulation in critically ill patients: a practical guideline. Ultrasound J. 2019;11(1):27. doi:10.1186/s13089-019-0144-5

7. Sou V, McManus C, Mifflin N, Frost SA. A clinical pathway for the management of difficult venous access. BMC Nurs. 2017;16(64):1–7. doi:10.1186/s12912-017-0261-z

8. Van Loon FHJ, Buise MP, Claassen JJF, Dierick-van Daele ATM, Bouwman ARA. Comparison of ultrasound guidance with palpation and direct visualisation for peripheral vein cannulation in adult patients: a systematic review and meta-analysis. Br J Anaesth. 2018;121(2):358–366. doi:10.1016/j.bja.2018.04.047

9. Egan G, Healy D, ÓNeill H, Clarke-Moloney M, Grace PA, Walsh SR. Ultrasound guidance for difficult peripheral venous access: systematic review and meta-analysis. Emerg Med J. 2013;30(7):521–526. doi:10.1136/emermed-2012-201652

10. Griffiths J, Carnegie A, Kendal R, Rajeev M. A randomised crossover study to compare the cross-sectional and longitudinal approaches to ultrasound-guided peripheral venepuncture in a model. Crit Ultrasound J. 2017;9(1):9. doi:10.1186/s13089-017-0064-1

11. Gabriel J. Understanding the challenges to vascular access in an ageing population. Br J Nurs. 2017;26(14):S15–S23. doi:10.12968/bjon.2017.26.14.S15

12. Dougherty L. Intravenous therapy in older patients. Nurs Stand. 2013;28(6):50–58. doi:10.7748/ns2013.10.28.6.50.e7333

13. Marsh N, Webster J, Mihala G, Rickard CM. Devices and dressings to secure peripheral venous catheters to prevent complications (Review). Cochrane Database Syst Rev. 2015;(6):CD011070. doi:10.1002/14651858.CD011070.pub2

14. Fields JM, Piela NE, Ku BS. Association between multiple IV attempts and perceived pain levels in the emergency department. J Vasc Access. 2014;15(6):514–518. doi:10.5301/jva.5000282

15. Gopalasingam N, Thomsen A-ME, Folkersen L, Juhl-Olsen P, Sloth E. A successful model to learn and implement ultrasound-guided venous catheterization in apheresis. J Clin Apher. 2017;32(6):437–443. doi:10.1002/jca.21533

16. Franco-Sadud R, Schnobrich D, Mathews BK, Candotti C. Recommendations on the use of ultrasound guidance for central and peripheral vascular access in adults: A position statement of the society of hospital medicine. J Hosp Med. 2019;14:E1–E22. doi:10.12788/jhm.3287

17. Ullman JI, Stoelting RK. Internal jugular vein location with the ultrasound Doppler blood flow detector. Anesth Analg. 1978;57(1):118. doi:10.1213/00000539-197801000-00024

18. Keyes LE, Frazee BW, Snoey ER, Simon BC, Christy D. Ultrasound-guided brachial and basilic vein cannulation in emergency department patients with difficult intravenous access. Ann Emerg Med. 1999;34(6):711–714. doi:10.1016/s0196-0644(99)70095-8

19. Clemmesen L, Knudsen L, Sloth E, Bendtsen T. Dynamic needle tip positioning – ultrasound guidance for peripheral vascular access. A randomized, controlled and blinded study in phantoms performed by ultrasound novices. Ultraschall Med. 2012;33(7):E321–E325. doi:10.1055/s-0032-1312824

20. Shamseer L, Moher D, Clarke M, Ghersi D. Preferred reporting items for systematic review and meta-analysis protocols (PRISMA-P) 2015: elaboration and explanation. BMJ. 2015;349:g7647. doi:10.1136/bmj.g7647

21. Bahl A, Pandurangadu AV, Tucker J, Bagan M. A randomized controlled trial assessing the use of ultrasound for nurse-performed IV placement in difficult access ED patients. Am J Emerg Med. 2016;34(10):1950–1954. doi:10.1016/j.ajem.2016.06.098

22. Bridey C, Thilly N, Lefevre T, Marie-Richard A. Ultrasound-guided versus landmark approach for peripheral intravenous access by critical care nurses; a randomised controlled study. BMJ Open. 2018;8:1-8. BMJ Open. 2018;8(6):e020220. doi:10.1136/bmjopen-2017-020220

23. Nishizawa T, Matsumoto T, Todaka T, Sasano M. Nurse-Performed Ultrasound-Guided Technique for Difficult Peripheral Intravenous Access in Critically Ill Patients: A Randomized Controlled Trial. J ASSOC VASC ACCESS. 2020;25(2):34–39. doi:10.2309/j.java.2020.002.001

24. McCarthy ML, Shokoohi H, Boniface KS, Eggelton R. Ultrasonography versus landmark for peripheral intravenous cannulation: a randomized controlled trial. Ann Emerg Med. 2016;68(1):10–18. doi:10.1016/j.annemergmed.2015.09.009

25. Söderström A, Nørgaard MS, Thomsen A-ME, Sørensen BS. Ultrasound-guidance of peripheral venous catheterization in apheresis minimizes the need for central venous catheters. Journal of Clinical Apheresis. 2020;35:200-205. J Clin Apher. 2020;35(3):200–205. doi:10.1002/jca.21780

26. Bauman M, Braude D, Crandall C. Ultrasound-guidance vs. standard technique in difficult vascular access patients by ED technicians. Am J Emerg Med. 2009;27(2):135–140. doi:10.1016/j.ajem.2008.02.005

27. Skulec R, Callerova J, Vojtisek P, Cerny V. Two different techniques of ultrasound-guided peripheral venous catheter placement versus the traditional approach in the pre-hospital emergency setting: a randomized study. Intern Emerg Med. 2020;15(2):303–310. doi:10.1007/s11739-019-02226-w

28. Costantino TG, Parikh AK, Satz WA, Fojtik JP. Ultrasonography-guided peripheral intravenous access versus traditional approaches in patients with difficult intravenous access. Ann Emerg Med. 2005;46(5):456–461. doi:10.1016/j.annemergmed.2004.12.026

29. Brannam L, Blaivas M, Lyon M, Flake M. Emergency nurses’ utilization of ultrasound guidance for placement of peripheral intravenous lines in difficult-access patients. Acad Emerg Med. 2004;11(12):1361–1363. doi:10.1197/j.aem.2004.08.027

30. Lamperti M, Bodenham AR, Pittiruti M, Blaivas M. International evidence-based recommendations on ultrasound-guided vascular access. Intensive Care Med. 2012;38(7):1105–1117. doi:10.1007/s00134-012-2597-x

31. Tran QK, Flanagan K, Fairchild M, Yardi I, Pourmand A. Nurses and Efficacy of Ultrasound-Guided Versus Traditional Venous Access: A Systematic Review and Meta-Analysis J Emerg Nurs. 2022;48(2):145-158.e1. doi:10.1016/j.jen.2021.12.003

